# Identification and validation of aging-related genes in IgA nephropathy

**DOI:** 10.1101/2025.02.20.25322599

**Authors:** Jinlian Shu, He Li, Hairong Li, Airong Xing, Shaofeng Yao

## Abstract

IgA nephropathy (IgAN) is the most common primary glomerulonephritis worldwide. Aging is a major risk factor for progression of IgAN to end stage renal disease. The purpose of this study was to identify and verify aging-related genes associated with IgAN through bioinformatics analysis. Microarray datasets of GSE93798 and GSE37460 were downloaded from the Gene Expression Omnibus (GEO) database. The aging-related DEGs (AR-DEGs) associated IgAN was analyzed by R programming software, and then genome ontology (GO) analysis and Kyoto Encyclopedia of Genes and Genomes (KEGG) analysis were performed. The PPI network of AR-DEGs was then contructed, and hub genes were ranked using five methods of the cytoHubba plugin in Cytoscape software. CIBERSORT algorithm was used to evaluate the infiltrating immune cells and their relationship with hub genes. Next, Nephroseq V5 online platform was used to verify and analyze the mRNA expression patterns of hub genes in IgAN patients and normal controls.A total of 372 DEGs were identified, of which the expression of 158 were upregulated and 214 were downregulated. GO and KEGG enrichment pathway analysis mainly focused on regulation of macrophage derived foam cell differentiation, PI3K-Akt signaling pathway. Based on the results of PPI network analysis, the eight hub genes were identified, including *AGT*, *ALB*, *CD36*, *EGF*, *KDR*, *LPL*, *MYC*, *PPARGC1A*. Immunoinfiltration analysis indicated that *CD36* were closely related to immune cell infiltration. Furthermore, the expression levels of these hub genes were validated using the Nephroseq V5 online platform. Further clinical sample studies confirmed that *CD36* was highly expressed in renal tissues of IgAN patients. These findings provide new insights into potential aging-related genes associated with IgAN, which may better contribute to understanding the pathogenesis of IgAN. *CD36* may have diagnostic value for aging-related IgAN.

## Introduction

IgA nephropathy (IgAN) is the most common primary glomerular disease worldwide. A biopsi-based study across multiple countries has shown that the overall population incidence of IgAN was at least 2.5 per 100,000[1]. Due to the possible unrecorded subclinical cases, the true prevalence and incidence of IgAN may be higher than the levels. In addition, the incidence of IgAN varies around the world. The prevalence of IgAN was significantly higher in Asian population, accounting for 40%-50% of primary glomerulonephritis in China[2]. IgAN patients were mostly young adults. However, with the increasing aging population, elderly IgAN has gradually become a great concern. An earlier study estimated that IgAN accounted for only 2.3% of all patients over the age of 50[3], but recent studies have reported that the proportion of elderly IgAN patients has increased to 18.3%[4]. These data suggested that the incidence of IgAN was increasing gradually in the elderly. Furthermore, studies had shown that 20%-40% of IgAN patients reached the endpoint of end stage renal disease within 20 years of diagnosis[5].

Aging, a process we all can’t avoid, is determined by a complex set of biological processes. Aging is the accumulation of aging cells with poor functions in the body, which leads to the reduction or loss of physiological functions, and ultimately leads to further exacerbation of functional decompensation. The main characteristics of aging are telomere attrition and genetic and epigenetic changes[6,7]. Many studies have shown that aging is a negative factor affecting the prognosis of IgAN[8,9]. A meta-analysis showed that the incidence of end-stage renal disease in elderly IgAN patients over 50 years was 1.95 times higher than that in non elderly IgAN patients[10]. A retrospective study investigating clinical and histopathological differences between elderly IgAN patients and middle-aged, younger-aged patients found that elderly patients had more comorbidity and higher blood pressure, lower estimated glomerular filtration rate (eGFR) and greater proteinuria at the time of kidney biopsy[11]. Histological analysis of older patients exhibit more chronic changes including interstitial fibrosis/tubule atrophy and arteriosclerosis. These results were consistent with other literature reports[12-14]. Above studies have showed that aging may be a key factor affecting the prognosis of IgAN patients, which may aggravate renal injury by promoting arteriosclerosis and interstitial fibrosis. However, whether aging-related genes play a critical role in the development of IgAN has remained unclear. Therefore, identifing the novel aging-related genes may provide new therpeutic targets and advance our understanding of pathogenesis in IgAN.

In addition, with regard to the pathogenesis of IgAN, the “four-hit” hypothesis is the most widely accepted, and accumulating evidence supported the involement of immune system in IgAN. T cells, B cells, monocytes and macrophages, and complement play crucial roles in the pathogenesis and progression of IgAN[15]. In addition, the results of single-cell RNA sequencing of peripheral blood mononuclear cells (PBMCs) showed that Th cells and B cells were more activated in IgAN patients, and the results of tissue biopsy staining demonstrated that the degree of renal CD4+T and B cell infiltration was positively correlated with the degree of renal damage[16]. Furthermore, previous studies have reported that immune senescence and chronic inflammation were the main biological changes in the aging process[17]. However, whether aging-associated immune response are related to aging-related IgAN remains unclear. In consequence, in order to further explore the potential molecular mechanisms and signaling pathways between IgAN and aging, we performed bioinformatics methods to comprehensively analyze aging-related genes and their association with immune infiltration in IgAN, which will provide potential therapeutic targets and biomarkers for the treatment of IgAN.

The rapid development and widespread application of high-throughput sequencing technology and numerous public databases can effectively screen disease-related biomarkers, providing important technical support for the prediction, diagnosis and treatment of human diseases. In this paper, we searched the gene expression profile of IgAN in the published Gene Expression Omnibus(GEO, https://www.ncbi.nlm.nih.gov/geo/).database, and then used GSE37460, GSE93798 as the object of this study. Aging related genes were collected by integrating GeneCards database (https://www.genecards.org/)[18] and MSigDB database (https://www.gsea-msigdb.org/gsea/msigdb)[19]. In this study, we used a variety of biological information analysis methods to screen aging-related IgAN genes, and verified them with Nephroseq V5 database and clinical specimens. This study design was depicted in Fig 1.

**Fig 1.**
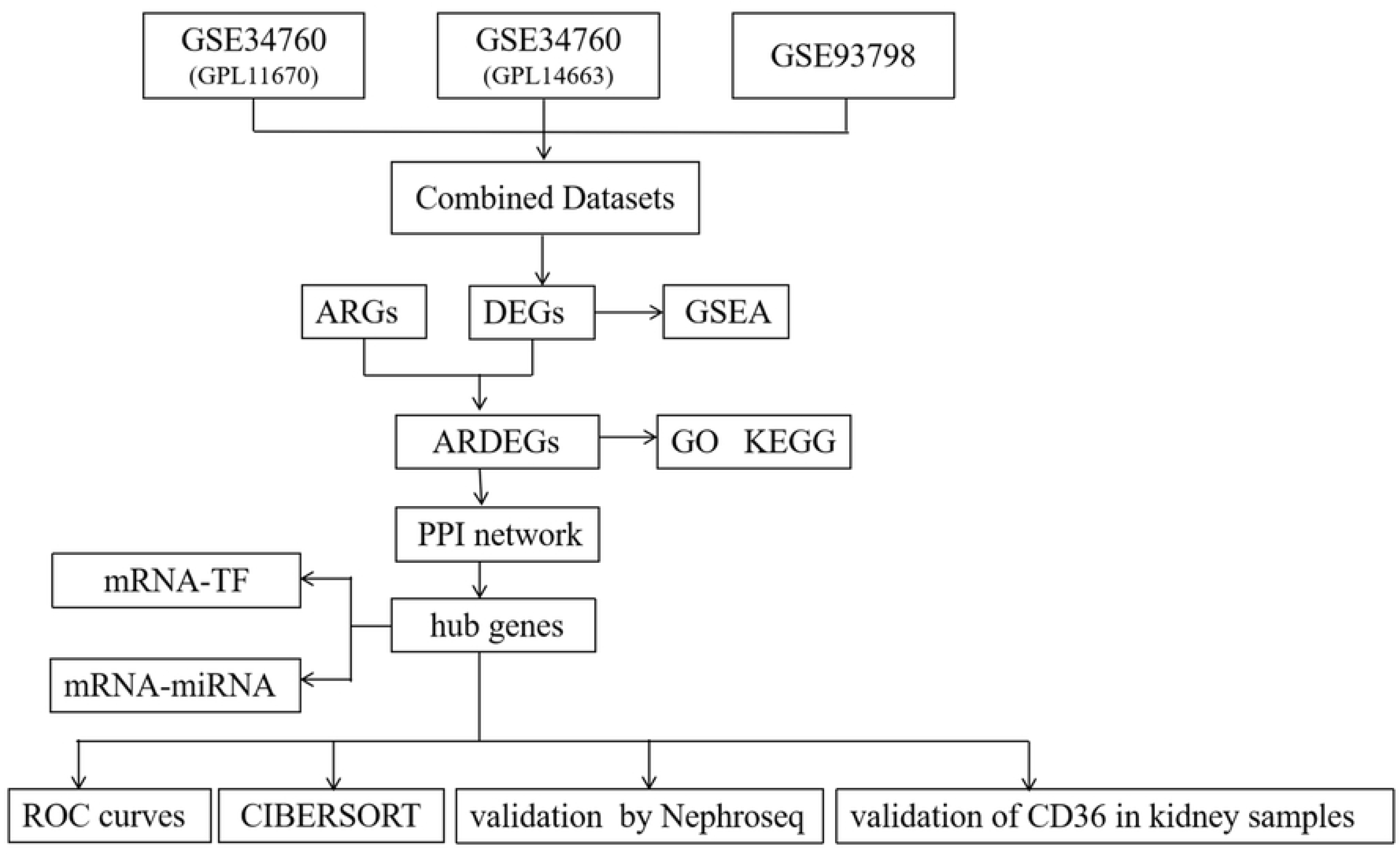
The design flow chart of the bioinformatics study. ARGs, aging-related genes; DEGs, differentially expressed genes; GSEA, Gene Set Enrichment Analysis; ARDEGs, Aging-related differentially expressed genes; GO, Gene Ontology; KEGG, Kyoto Encyclopedia of Genes and Genomes; PPI network, Protein-protein Interaction Network; TF, Transcription Factor; ROC, Receiver Operating Characteristic Curve.

## Methods

### Microarray data collection

We downloaded the IgAN dataset GSE374603[20], GSE937984[21] from the GEO database using the R package GEOquery. The samples of datasets GSE37460 and GSE93798 were all taken from human renal biopsy specimens. GSE37460 datasets was based on GPL11670 (Affymetrix Human Genome U133 Plus 2.0 Array [Hs133P_Hs_ENTREZG.cdf]) platform and GPL14663 (Affymetrix GeneChip Human Genome HG-U133A Custom CDF [Affy_HGU133A_CDF_ENTREZG_10]) platform. Among them, the GPL11670 platform data in dataset GSE37460 contained 18 healthy control samples; The GPL14663 platform data in dataset GSE37460 included 27 IgAN samples and 9 healthy control samples. 20 IgAN samples and 22 control samples were contained in the GSE937984 dataset which was based on GPL22945 ([HG-U133_Plus_2] Affymetrix Human Genome U133 Plus 2.0 Array [CDF: Brainarray HGU133Plus2_Hs_ENTREZG_v19]) platform. The specific information was shown in S1 Table.

The GeneCards database and the MSigDB database were used to collect aging-related genes (ARGs). The GeneCards database is a searchable and comprehensive database containing almost all known human genes. We searched GeneCards database for the aging-related genes by searching “aging” as keywords, obtaining 259 ARGs and retaining only the genes with "Protein Coding" and "Relevance Score > 12". Similarly, we downloaded 110 aging-related genes from the MSigDB database by searching the keyword "aging". Subsequently, a total of 355 ARGs were finally obtained after merging and elimination of duplicates. The detailed information was shown in S1 Table.

The datasets GSE37460 and GSE93798 were processed through the "SVA" R package to remove the batch effect and then obtained the combined GEO datasets. The combined datasets included 47 IgAN samples and 49 control samples. Finally, the combined GEO datasets were standardized and normailiezd with probe annotation by usin "limma" package of R software. To test for batch effects, principal component analysis (PCA) of the expression matrix was performed before and after removal of batch effects. PCA is a method of dimensionality reduction of data by converting high-dimensional data to low-dimensional data and displaying these features in a two-dimensional or three-dimensional graphs.

### Identification of aging-related differentially expressed genes in IgAN

The differentially expressed genes (DEGs) between IgAN and control group in combined GEO datasets were selected using the limma R package. The cut-off criteria was set as adjusted P-value < 0.05 and |logFC| > 0.5 in the combined datasets. P values were adjusted using the Benjamini-Hochberg (BH) method. The volcano plots for the DEGs expression were plotted using R package ggplot2. Subsequently, we used R package to visually draw the venn diagram of ARGs and DEGs, thus identifying the differentially expressed aging-related genes(AR-DEGs) associated IgAN. Heat map of AR-DEGs was shown with R package pheatmap and chromosome localization map was drawn by R package RCircos.

### Gene Set Enrichment Analysis (GSEA)

Gene Set Enrichment Analysis (GSEA) was a computational method which was used to evaluate the strength of gene-phenotype correlation within a given gene set[22]. In this study, the genes in the combined GEO datasets were first sequenced according to logFC values, and then the R-package clusterProfiler was applied to implement GSEA. The parameters used in the GSEA were as follows: the seed is 2022, the number of computations is 1000, and the minimum number of genes contained in each gene set is 10 and the maximum number of genes is 500. GSEA was performed using the c2 gene sets. Cp. all. V2022.1. Hs. symbols gene sets which were obtained from the MSigDB database. Adj.p < 0.05 and FDR q value < 0.05 were regarded as statistically significant. The p value was corrected using BH method.

### GO and KEGG enrichment Analysis

To further evaluate the biological functions and signaling pathways of AR-DEGs, we carried out Gene Ontology (GO) and The Kyoto Encyclopedia of Genes and Genomes (KEGG) analyses by clusterprofiler R package, with adj.p < 0.05 and false discovery rate (FDR) q value < 0.05 as the threshold. GO enrichment analysis included three aspects: molecular function (MF), biological process (BP) and cellular component (CC). The p value was corrected using BH method.

### Protein-Protein Interaction (PPI) Network Construction and identification of hub genes

In order to further assess the functional interactions among AR-DEGs, a PPI network was constructed using the Search Tool for the Retrieval of Interacting Genes (STRING)(http://string-db.org/)[23].The minimum correlation coefficient greater than 0.700 was considered to be significant. The PPI network was then visualized using Cytoscape and hub genes were identified by cytoHubba plugin. Among them, the following five algorithms are used in Cytohubba plugin, namely maximal clique centrality(MCC), closeness, maximum neighborhood component(MNC), degree, edge percolated component(EPC). In the PPI network, the scores of AR-DEGs were computed and then the top 10 genes were selected. The intersection of genes obtained by these five algorithms were identified as the hub genes and visually displayed by generating venn diagrams.

### Construction of regulatory network

Transcription factors (TF) control gene expression through interaction with hub genes during the post-transcriptional stage. To further explore the relationship between TF and the hub genes, we applied ChIPBase database (http://rna.sysu.edu.cn/chipbase/) to predict TF targeted by hub genes[24]. In addition, the miRNA-TF interaction pairs were screened using the sum of Number of samples found (upstream) and Number of samples found (downstream) greater than 4 as thresholds. Then, the mRNA-TF regulatory network was visualized by cytoscape software.

MiRNA plays an important regulatory role in the process of biological processes. A single miRNA can regulate multiple target genes, while multiple miRNAs can target a single gene. To better understand the correlation between hub genes and miRNA, we used the ENCORI database to analysis mRNA-miRNA interaction[25]. PancancerNum > 8 was regarded as the screening criteria for mRNA-miRNA. Then, the mRNA-miRNA co-regulation network was visualized through Cytoscape.

### Differential expression verification and ROC curve analysis of hub genes

To further verify hub genes, we used R language to analyze the differences in hub genes expression between IgAN group and control group. Differences in the expression of hub genes between IgAN group and control group were compared using Mann-Whitney test (Wilcoxon rank sum test) in combined datasets. The area under the curve (AUC) of the ROC curve was used to validate the diagnostic efficiency of IgAN using the "pROC" R package based on hub genes.

### Immune infiltration analysis

CIBERSORT is based on the principle of linear support vector regression to deconvolute the transcriptome expression matrix to estimate the composition and abundance of immune cells in the mixed cells[26]. The results of immune cell infiltration matrix in the combined datasets were finally obtained through CIBERSORT combined with LM22 signature matrix. Subsequently, immune cells with significant differences between IgAN groups and control groups were screened for subsequent analysis. The correlation between hub genes and immune cells was computed using Spearman’s correlation, and bubble plots were drawn in R using the ggplot2 package.

### Nephroseq V5 Validation

In order to verify the expression level of hub genes in IgAN, we used Nephroseq V5 online platform (http://v5.nephroseq.org) to analyze hub genes[27]. Group data were represented as mean ± standard deviation. Student’s t test were used to compare the differences between two groups. Statistical significance was shown as *p < 0.05,**p < 0.01,***p < 0.001.

### Immunohistochemistry staining

The clinical renal tissue sample validation study was conducted in accordance with the Helsinki Declaration with the consent of the Ethics Committee of the Second People’s Hospital of Hefei. The kidney biopsy specimens were collected by percutaneous renal puncture biopsy technique in the department of nephrology from January 2021 to May 2024. The kidney tissue samples were prepared according to standardized methods and analyzed by light microscopy, electron microscopy and immunofluorescence microscopy respectively in the department of pathology. A total of 22 patients with IgAN were included as the case group, and 11 samples of healthy kidney tissues which were taken after renal tumor operation were selected as the control group. The inclusion criteria for IgAN were (1) age ≥ 18 years; (2) No corticosteroids and immunosuppressive therapy were used within one month prior to enrollment; (3) CKD stages 1-4, eGFR was calculated using CKD-EPI formula. Exclusion criteria: (1) diagnosis of secondary IgAN, including liver disease, anaphylactoid purpura, ulcerative colitis, systemic lupus erythematosus, tumors, etc.; (2) Combined with infection, malignant hypertension, cardiac insufficiency, etc.; (3) The follow-up data were incomplete. The biochemical data for patients are summarized in Table 1.

**Table 1.**
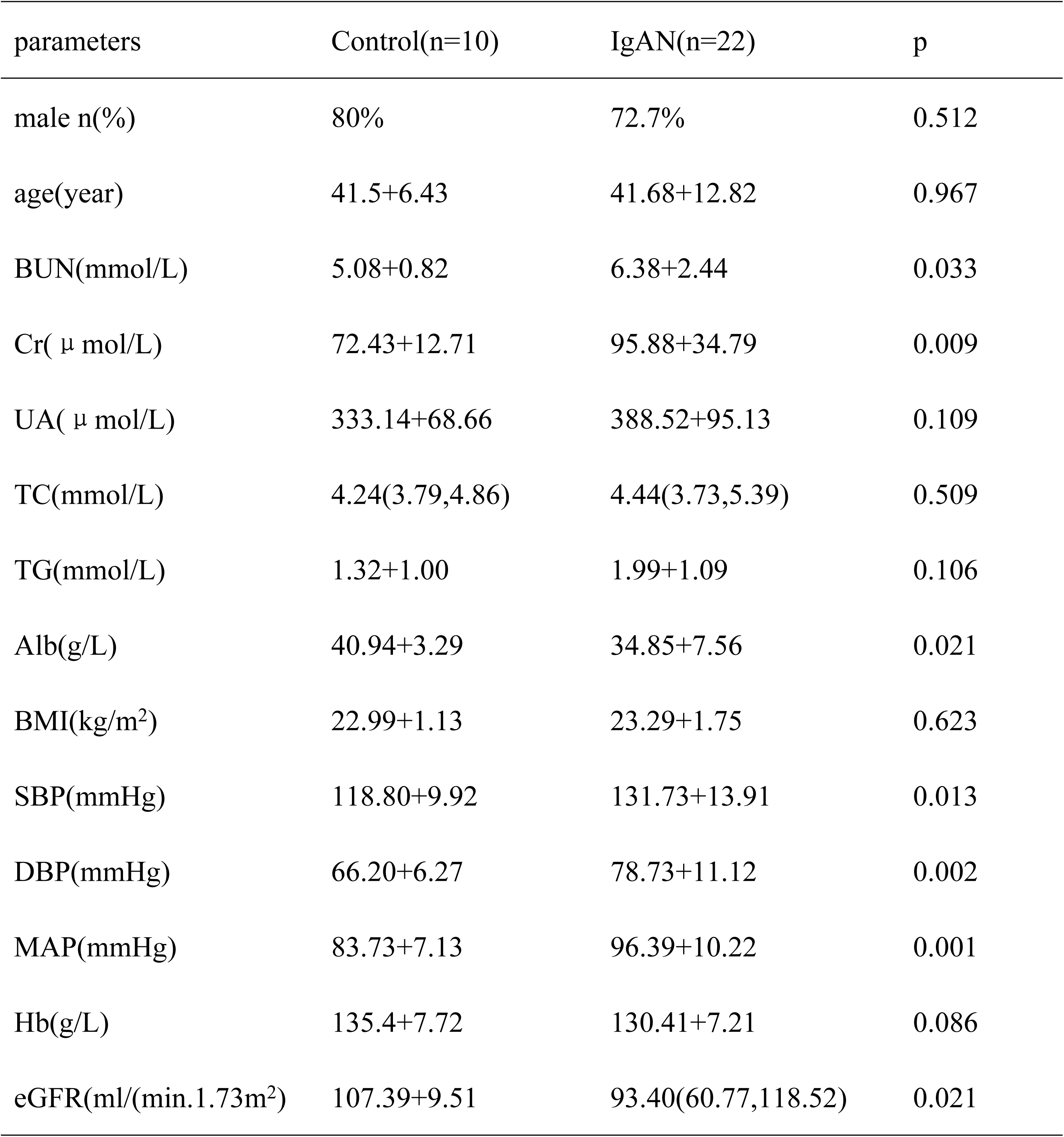
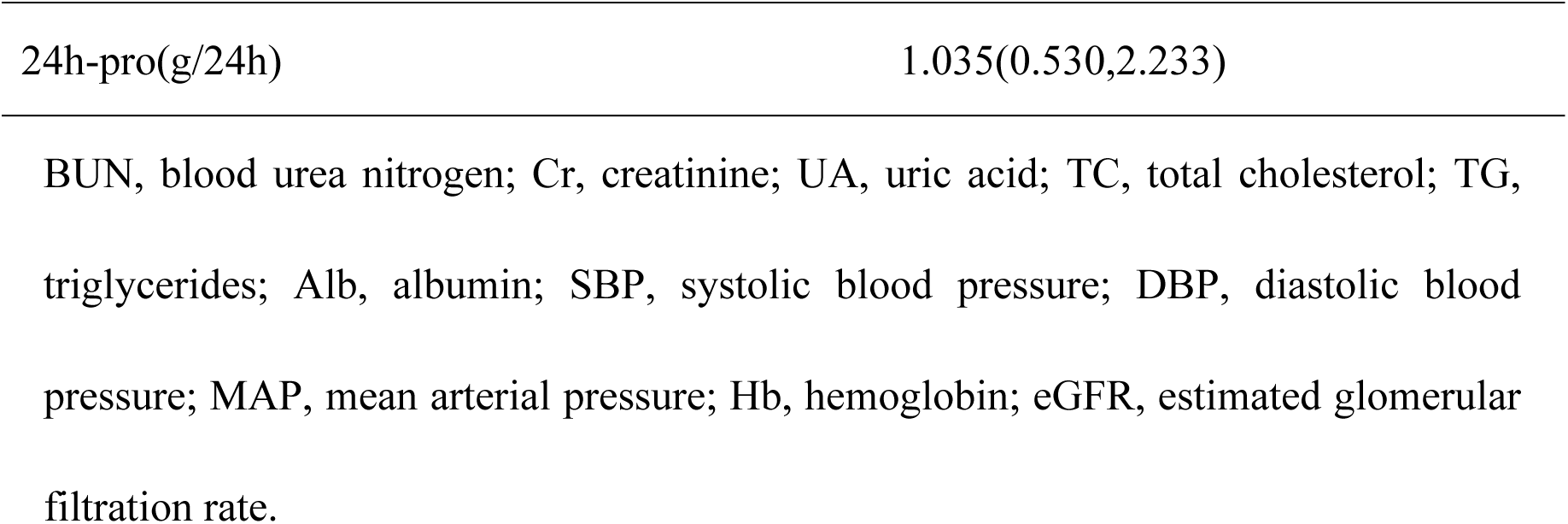
Clinical characterisitics.

Paraffin-embedded kidney sections were dewaxed in xylene and hydrated in ethanol. Antigen retrieval was performed on tissue samples with EDTA buffer (pH 9.0) in a pressure cooker for 2 min. Subsequently, the slices were then incubated at room temperature in 3% hydrogen peroxide for 10 minutes to block endogenous peroxidase activity, rinsed three times with distilled water followed by PBS-T. After washing, the sections were capped and incubated with the primary anti-CD36 antibody(Bioss, 1:300) for 60min at 37℃. Following the antibody incubation, the slices were removed from the incubator and rinsed with PBS-T for 3 times. Then, the extra liquid was blotted off and the slices were incubated with HRP-conjugated secondary antibodies at 37℃ for 30min. Subsequently, the slices were removed again and rinsed 3 times with PBS-T. Removal of excess liquid, the slices were incubated in the DAB solution until developed brown color. The sections were then rinsed with distilled water and counterstained with hematoxylin, rendered transparent by xylene. Finally, the results were observed under an optical microscope after the sections were sealed with neutral gum. Five random visual fields were selected for per section, and then the positively stained areas were calculated using Image J software.

### Statistical analysis

Statistical analysis was performed using R (version 4.3.1) and SPSS (version 20.0), while protein expression levels in immunohistochemical images were quantified using ImageJ (version 2.3.0). Measurement data that meet the criteria of normality and homogeneity of variance were expressed as mean ± standard deviation (x ± s); otherwise, they were presented as median (M) and interquartile range (p25, p75). When the assumptions of normality and homogeneity of variance are satisfied, the t-test was used to compare the means of two groups. For non-normally distributed variables, the Mann-Whitney U test was applied. Categorical data were expressed as percentages, and comparisons between percentages were performed using the chi-square (χ^2^) test. p value < 0.05 was considered statistically significant.

## RESULTS

### Identification of DEGs and AR-DEGs in IgAN

First, to obtain DEGs related IgAN, we used R-packet sva to eliminated batch effects from the gene expression matrix after merging the GSE104954 and GSE30528 datasets. The box diagram (Fig 2A-B) clearly presented that the difference in the expression values of the datasets before and after batch impact removal. In addition, the principal component analysis also showed that the distributioin of datasets before and after removing batch effects(Fig 2C-D). The above results showed that the sample distributions among IgAN datasets were consistent after removing the batch impact. After the microarray results were normalized, totally 372 DEGs involved in IgAN were identified by limma package (adj.p < 0.05, |logFC| > 0.5), of which 158 genes were up-regulated and 214 genes were down-regulated. The volcano plot was represented in Figure 3A. Subsequently, 20 AR-DEGs were obtained by intersection of DEGs involved in IgAN with 355 aging-related genes, namely *ABCB1, AGT, AGTR1, ALB, ARG2, C3, CD36, CDKN1A, COL1A1, CX3CR1, EG, GHR, HBB, HLA-DQB1, KDR, LPL, MYC, PPARGC1A, SERPINE1, TWIST1*. A vene diagram were shown in Figure 3B. Heat map of AR-DEGs in the IgAN groups and control groups was displayed using R package pheatmap(Fig 3C). R-package RCircos was used to draw a chromosome localization map of AR-DEGs. The chromosome localization diagram showed that more genes were located on the 4 and 7 chromosome(Fig 3D). *ALB, EGF, KDR, PPARGC1A* were located on chromosome 4, whereas *ABCB1, CD36, SERPINE1, TWIST1* were located on chromosome 7.

**Fig 2.**
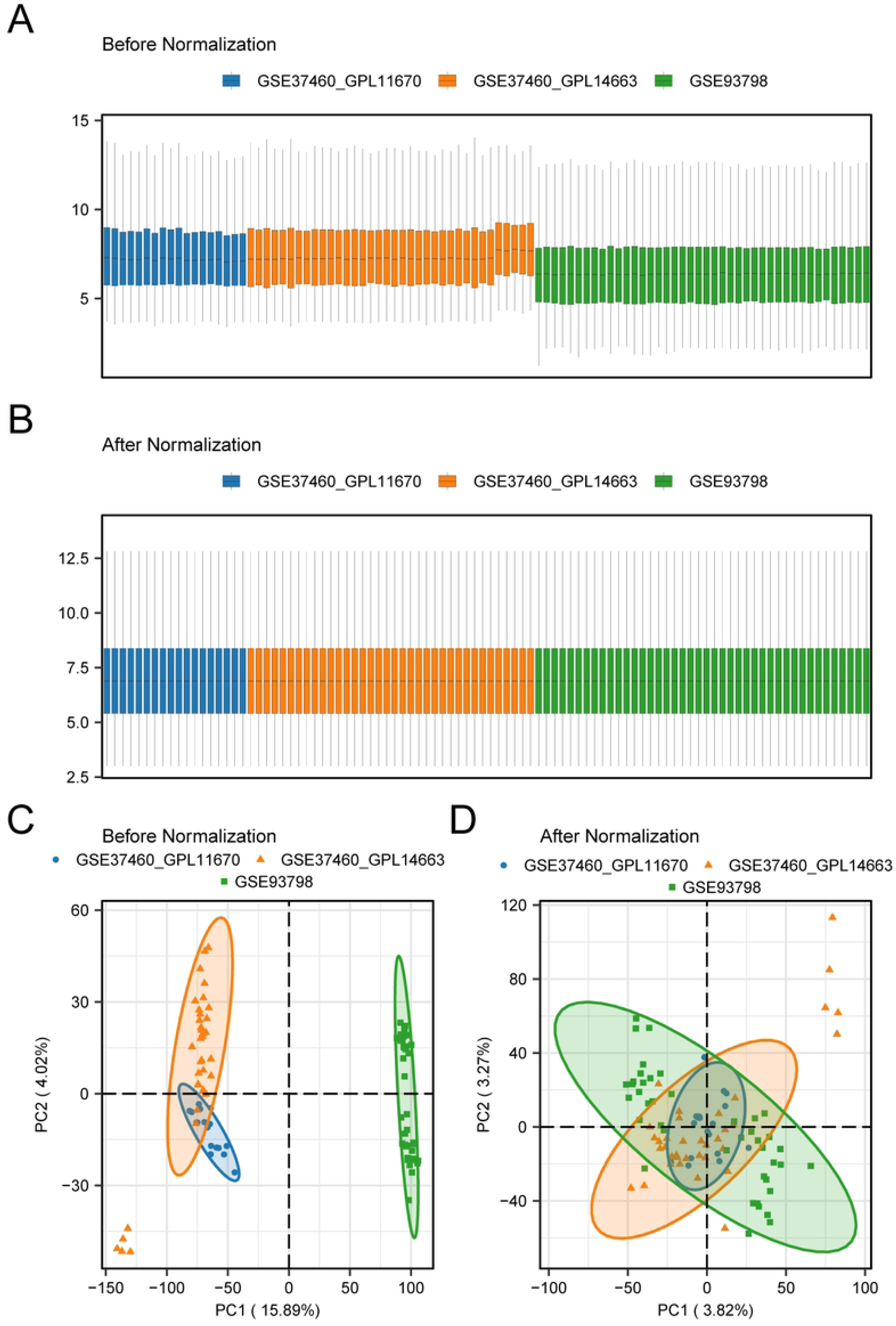
GEO datasets de-batching. A: Box plot of the datasets before de-batching. B: Gene expression levels statistics of the integrated dataset after de-batching. C: Principal component analysis (PCA) between datasets before de-batching. D: PCA of the integrated datasets after de-batching.

**Fig 3.**
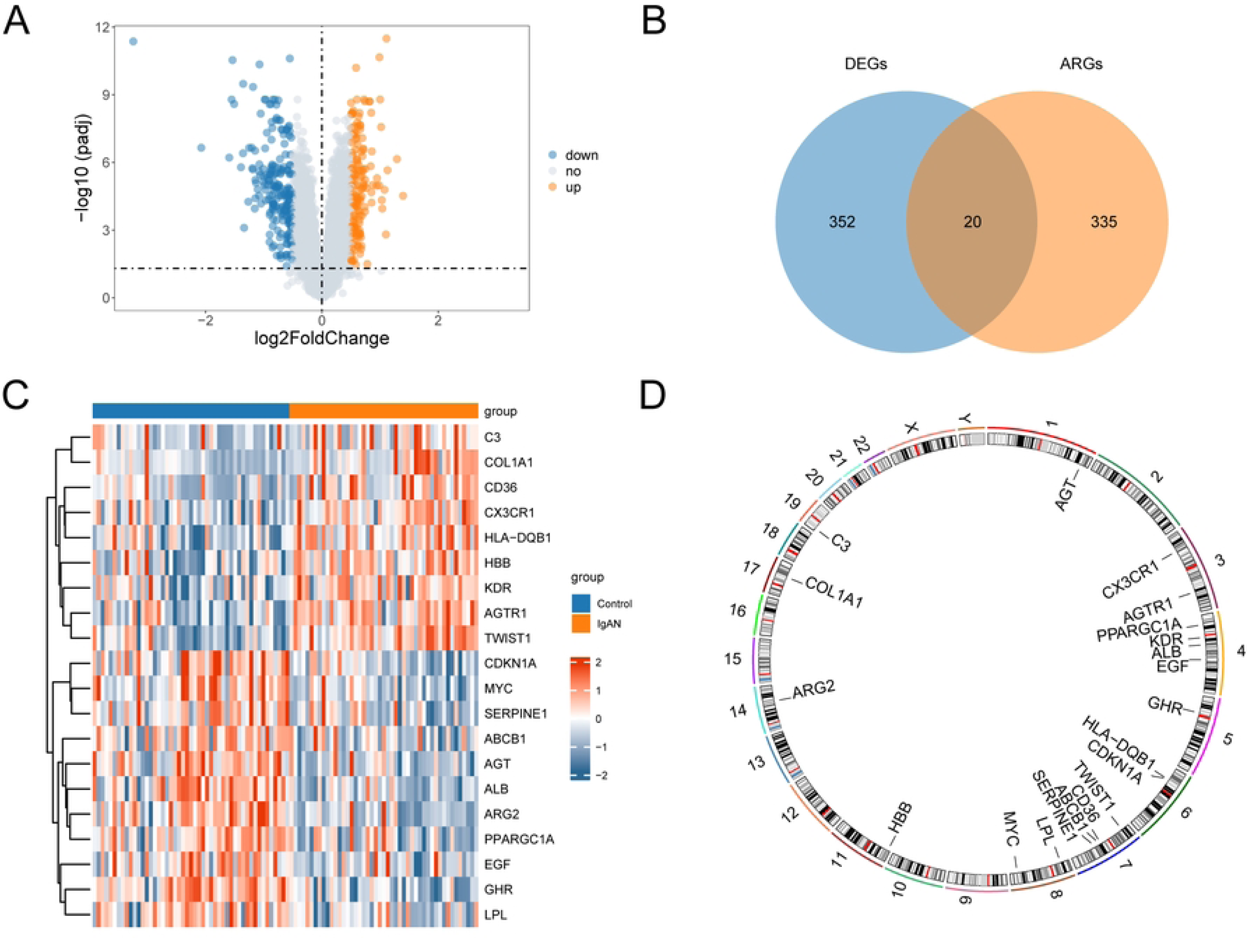
Identification of Aging-related genes (ARGs) and differentially expressed aging-related genes (AR-DEGs). A: Volcano plot of differentially expressed genes (DEGs) between IgAN group and control group in the integrated GEO Datasets. The red nodes represent upregulated genes, the blue nodes represent downregulated genes and the gray nodes represent nonsignificant genes; B: Venn diagram of ARGs and DEGs. Blue represents DEGs, pink represents ARGs; C: Heat map of ARDEGs in the integrated GEO Datasets: orange indicates IgA nephropathy samples, blue indicates normal control samples, red represents high gene expression, and blue represents low gene expression; D: Chromosomal mapping of ARDEGs.

### Gene Set Enrichment Analysis (GSEA)

We performed GSEA to explore the potential functions and the key pathways in IgAN(Fig 4A). The results suggested that most of the enriched gene sets were significantly enriched in reactome assembly of collagen fibrils and other multimeric structures (Fig 4B), reactome signaling by notch4 (Fig 4C), reactomen negative regulation of notch4 signaling (Fig 4D), reactome costimulation by the CD28 family (Fig 4E), reactome met promotes cell motility (Fig 4F), wp inflammatory resoponse pathway (Fig 4G), reactome hedgehog ligand biogenesis (Fig 4H) and other biologically relevant functions and signaling pathways. Specific details of the results are summarized in S2 table.

**Fig 4.**
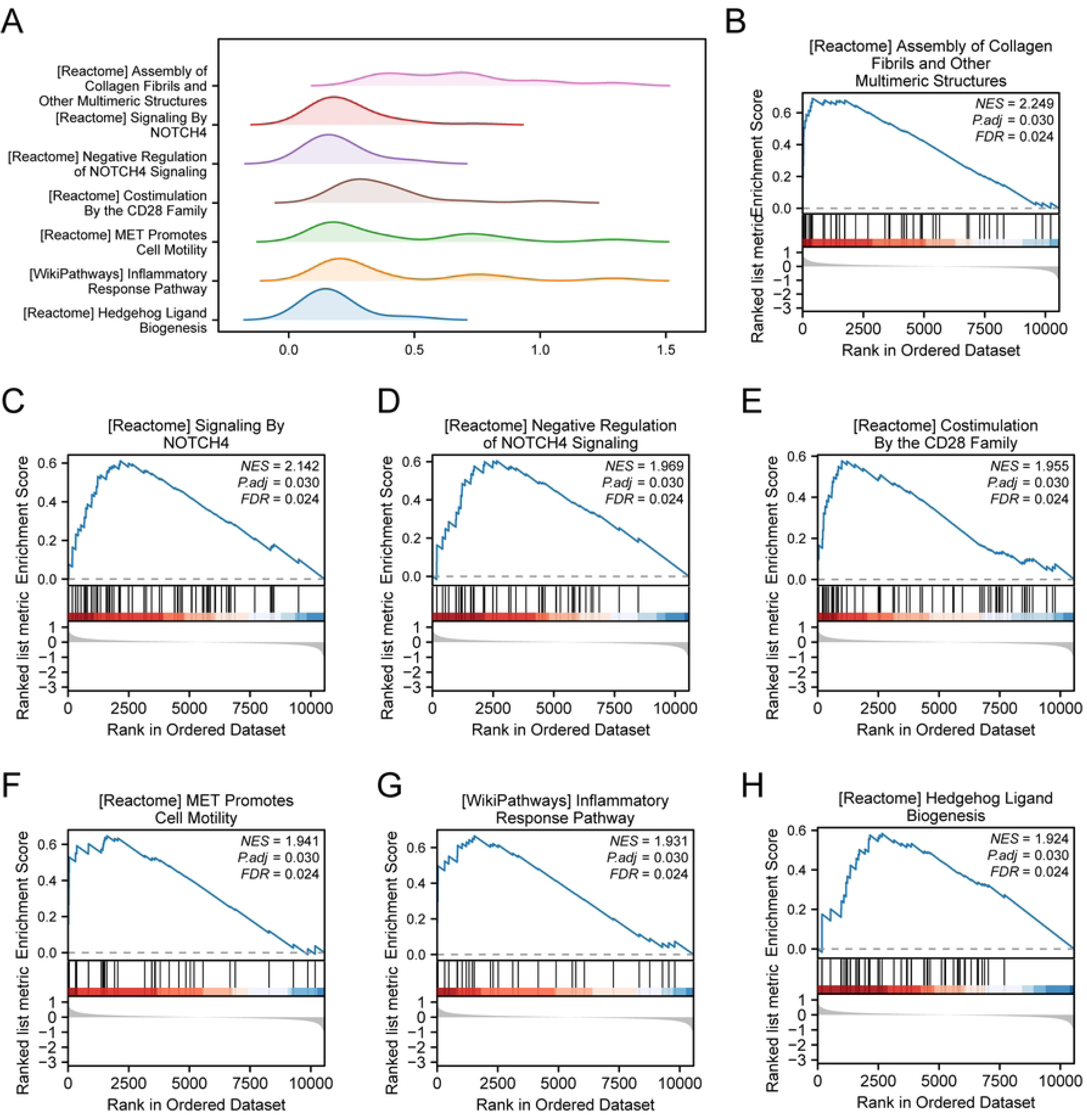
The results of Gene set enrichment analysis (GSEA). A: GSEA method was performed to rank the effects of differentially expressed biological pathways. B: GSEA results of Reactome assembly of collagen fibrils and other multimeric structures chain. C: GSEA results for Reactome signaling by NOTCH4. D: GSEA results of Reactome negative regulation of NOTCH4 signaling chain. E: GSEA results for Reactome costimulation by the CD28 family. F: GSEA results of Reactome MET promotes cell motility chain. G: GSEA results for the wikipathway inflammatory response pathway. H: GSEA results for Reactome hedgehog ligand biogenesis. NES, Normalized Enrichment Score; FDR, False Discovery Rate.

### Go and KEGG enrichment analysis of AR-DEGs

In order to further assess the potential biological signature of these AR-DEGs, we then performed GO and KEGG enrichment analysis by using R software. As shown in the S3 Table, the GO analysis showed that AR-DEGs were significantly enriched in biological processes (BP), including positive regulation of macrophage derived foam cell differentiation, nitric oxide biosynthetic process, nitric oxide metabolic process, reactive nitrogen species metabolic process, regulation of macrophage derived foam cell differentiation, macrophage derived foam cell differentiation. GO molecular function (MF) were notably concentrated in growth factor binding, G protein-coupled receptor binding, cytokine binding, lipoprotein particle binding, protein-lipid complex binding. Additionally, GO cellular component (CC) analysis revealed that these genes were mainly focused on platelet alpha granule, blood microparticle, platelet alpha granule lumen, external side of plasma membrane, secretory granule lumen. Furthermore, KEGG pathway analyses exhibited that AR-DEGs were mostly enriched in AGE-RAGE signaling pathway in diabetic complications, proteoglycans in cancer, bladder cancer, PI3K-Akt signaling pathway, endometrial cancer. Figure 5A was the GO/KEGG visual histogram. Meanwhile, the network diagram of biological process (BP), cellular component (CC), molecular function (MF) and biological pathway (KEGG) was drawn according to GO and KEGG analysis (Fig 5B-E).

**Fig 5.**
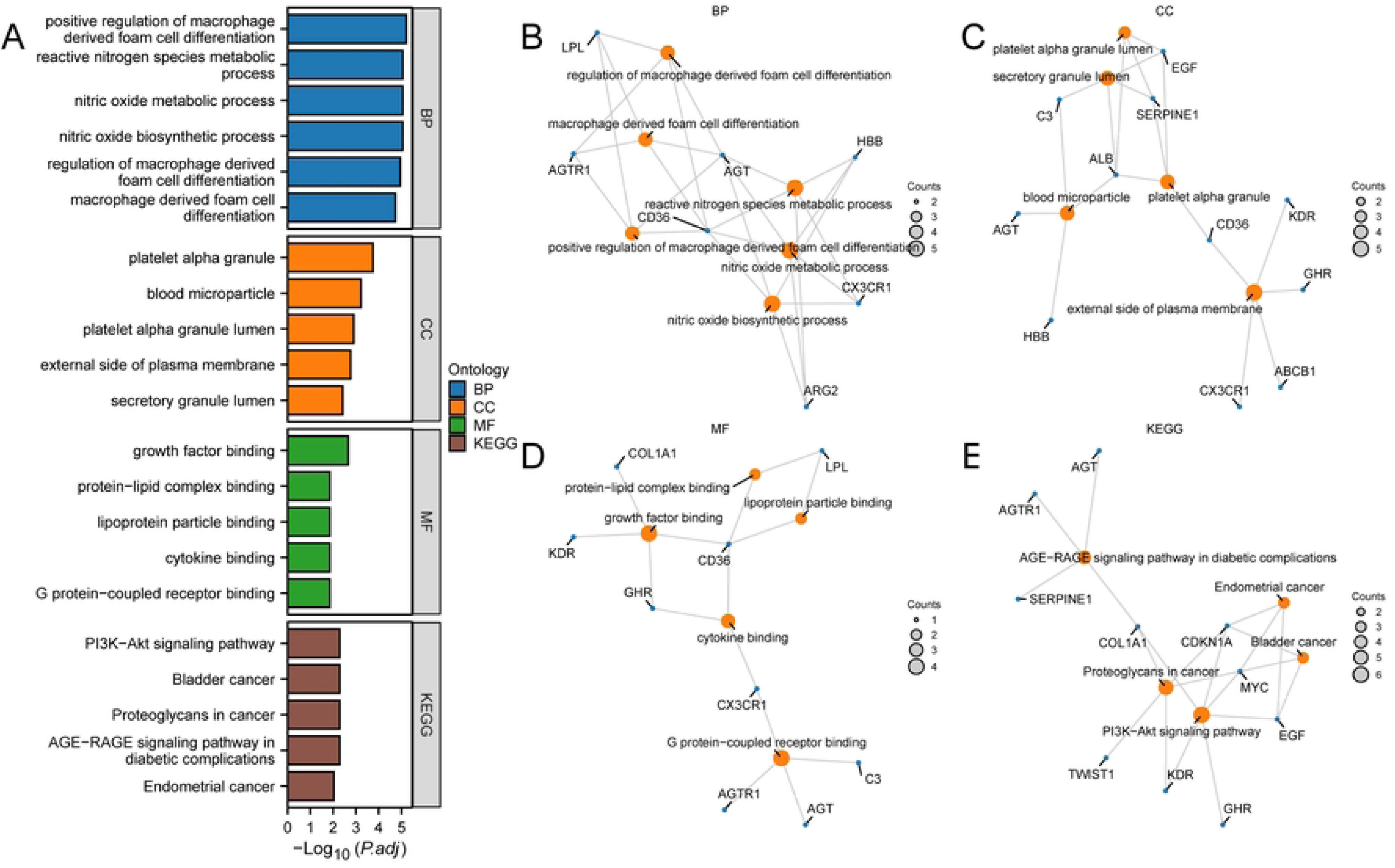
Gene Ontology (GO) and Kyoto Encyclopedia of Genes and Genomes (KEGG) Enrichment Analysis for ARDEGs. A: Bar graph of GO and KEGG enrichment analysis results of ARDEGs. B: GO and KEGG enrichment analysis results of ARDEGs were shown in the network diagram: BP (B), CC (C), MF (D) and KEGG (E). The orange-red nodes represent items, the blue nodes represent molecules, and the lines represent the relationship between items and molecules. BP, biological process; CC, cellular component; MF, molecular function.

### Construction of PPI network and identification of hub genes

To investigate the interaction relationship among these AR-DEGs, a PPI network was constructed and analyzed using STRING database (Fig 6A). The network showed that there were links between 15 genes, namely: *AGT, AGTR1, ALB, C3, CD36, CDKN1A, COL1A1, EGF, GHR, KDR, LPL, MYC, PPARGC1A, SERPINE1, TWIST1*. Figure 6B-F shows PPI networks mapped by the top 10 AR-DEGs of the previously mentioned five algorithms. The intersection of the top 10 genes which ranked by previous mentioned methods were hub genes. The hub genes were *AGT, ALB, CD36, EGF, KDR, LPL, MYC, PPARGC1A* respectively (Fig 6G).

**Fig 6.**
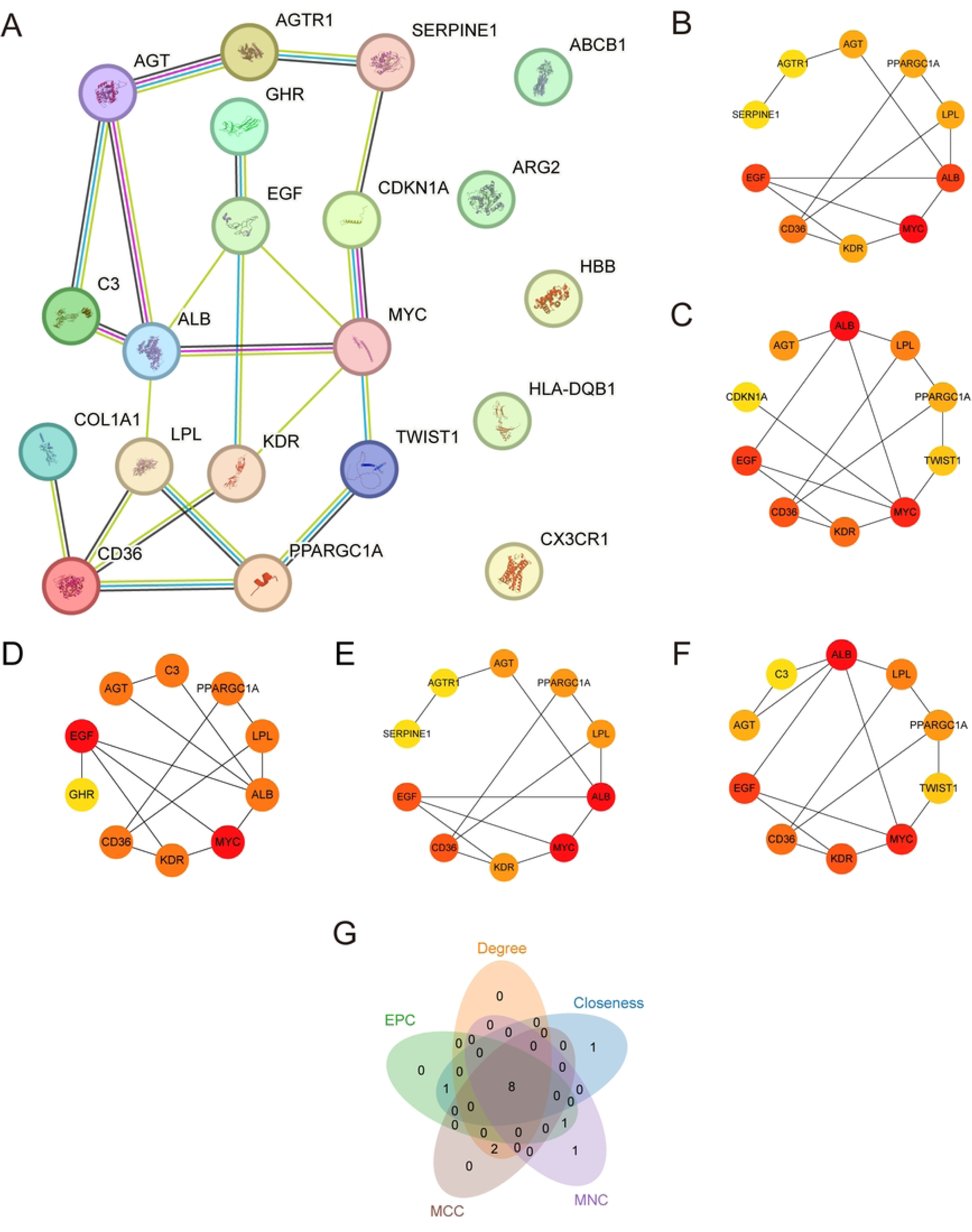
PPI Network and hub genes analysis. A: The PPI Network of ARDEGs calculated from STRING database. The PPI Network of the TOP10 ARDEGs calculated by the 5 algorithms of the CytoHubba plugin, including MCC (B), Closeness (C), MNC (D), Degree (E) and EPC (F). G: The Venn diagram of top 10 ARDEGs by the 5 algorithms of the CytoHubba plugin.

### Construction of regulatory network

The corresponding TF were obtained by ChIPBase database based on the hub genes, the mRNA-TF regulatory network was constructed and visualized using cytoscape software, which consisted of 7 nodes and 36 edges(Fig 7A).

**Fig 7.**
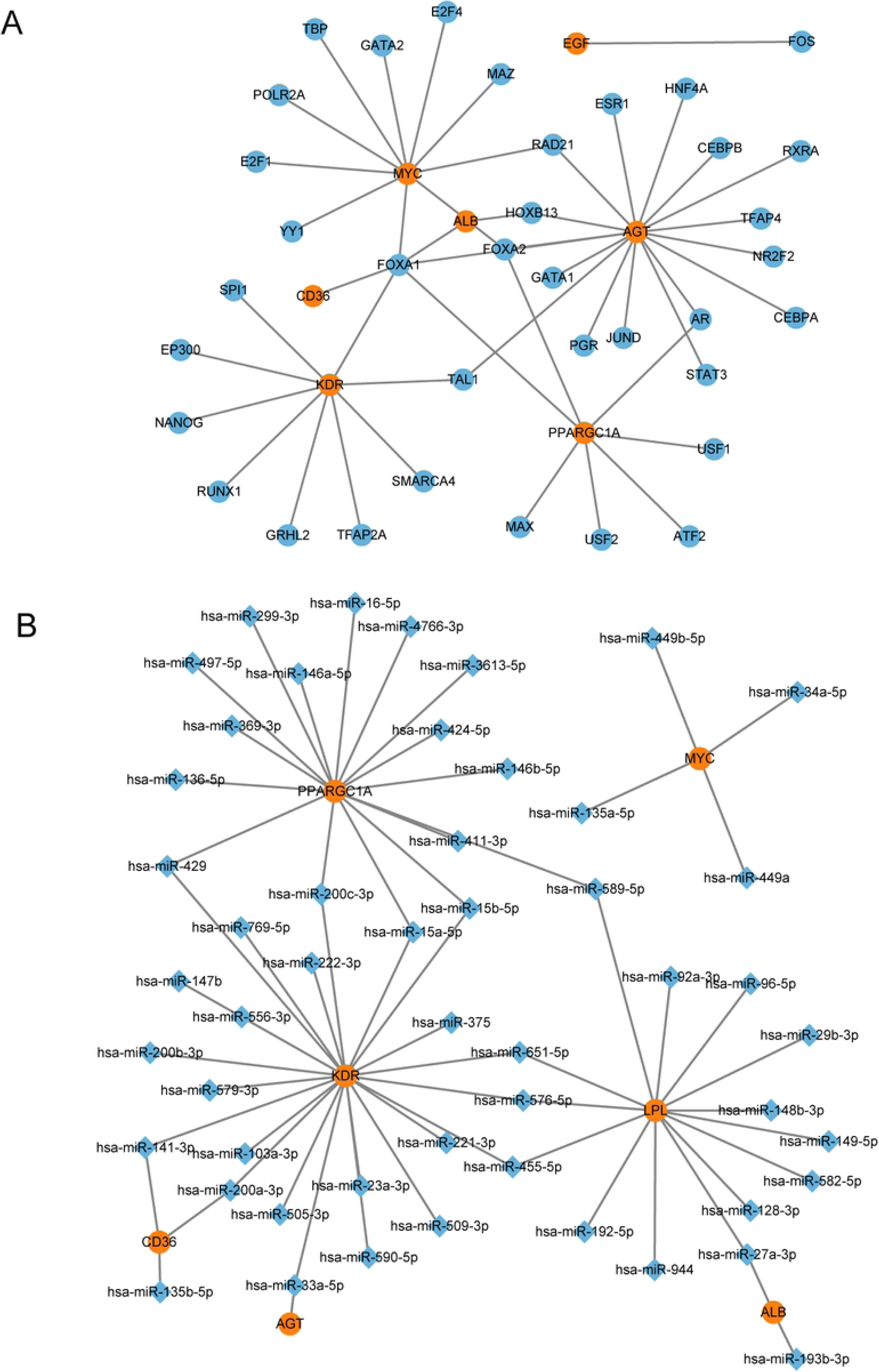
Regulatory Network of hub genes. A: Network for TF-gene interaction with hub genes. B: Gene-miRNA co-regulation network. The organge color nodes indicate hub genes, the blue nodes indicate TF, and the blue diamonds for miRNA. TF, Transcription Factor;

Based on these eight AR-DEGs, mRNA-miRNA co-regulatory network was constructed by ENCORI database, and the results showed that the network contained 7 nodes and 51 edges(Fig 7B).

### Expression level analysis and ROC curve analysis of hub genes

Compared with the control groups, the expression levels of CD36 and KDR in IgAN group were significantly up-regulated, while the expression levels of *AGT, ALB, EGF, LPL, MYC, PPARGC1A* were significantly decreased in IgAN group (p value < 0.001)(Fig 8A).

**Fig 8.**
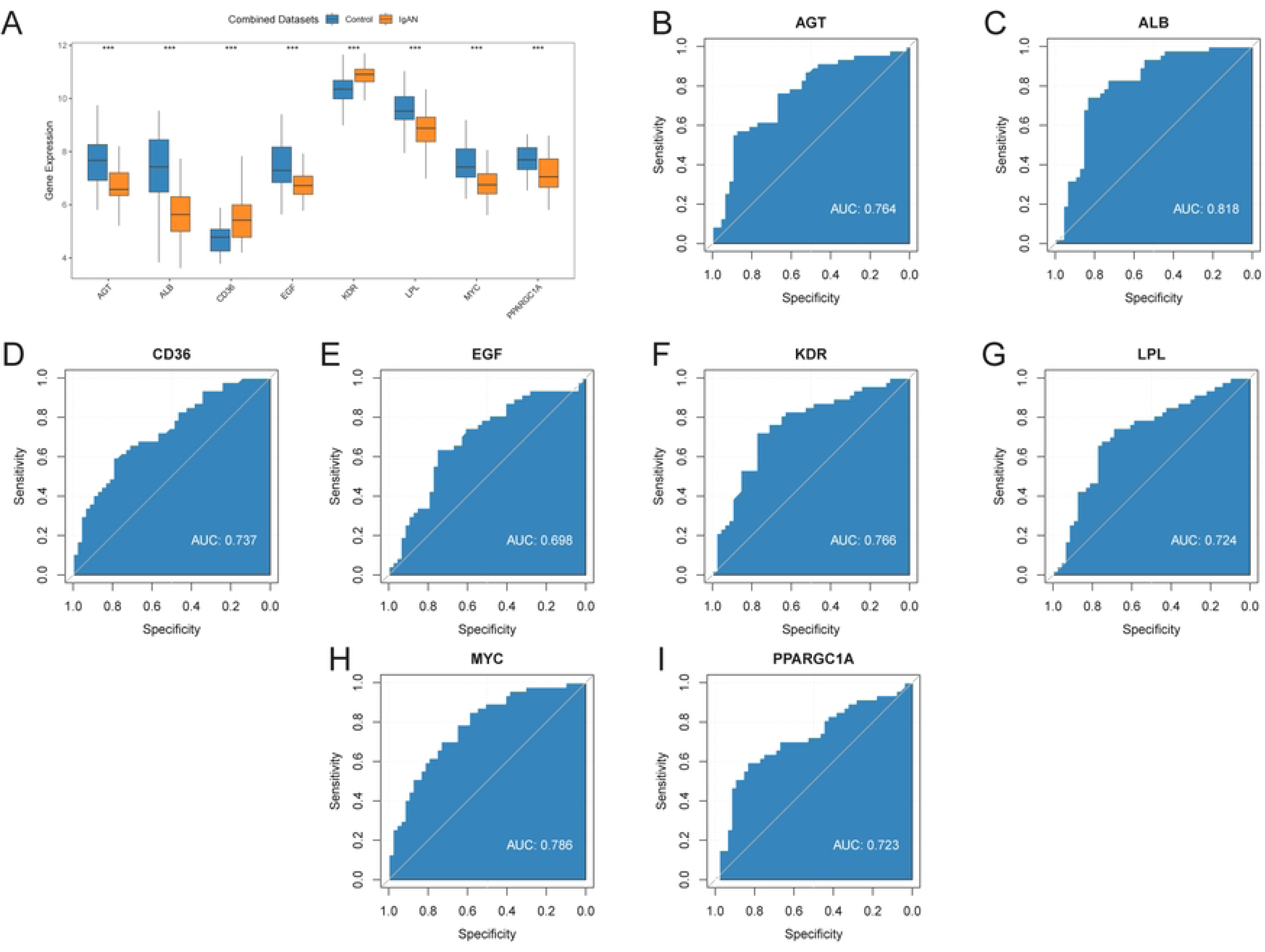
Differential Expression Validation and ROC Curve Analysis. A: the expression levels of hub genes between IgAN and controls. ROC curves of *AGT* (B), *ALB* (C), *CD36* (D), *EGF* (E), *KDR* (F), *LPL* (G), *MYC* (H) and *PPARGC1A* (I) in IgAN. ROC, Receiver Operating Characteristic Curve.

Based on the above analysis, we drew ROC curves and calculated the AUC to validate the diagnostic value of hub genes in the combined datasets. The AUC for *AGT, ALB, CD36, EGF, KDR, LPL, MYC, PPARGC1A* in IgAN patients and healthy control were 0.764, 0.818, 0.737, 0.698, 0.766, 0.724, 0.786, 0.723 respectively(Fig 8B-I).

### Immune Infiltration Analysis (CIBERSORT)

The relative abundance of 22 immune cells in IgAN were estimated by the CIBERSORT algorithm. As shown in Figure 9A, the proportion of 22 immune cells was plotted as a histogram. The expression levels of five kinds of immune cells were statistically increased in IgAN group compared with control group, namely macrophages M1, macrophages M2, monocytes, NK cells activated, T cells CD8, whereas B cells naive, neutrophils, NK cells resting, T cells CD4 memory resting were decreased in IgAN group(Fig 9B). The heat map presented the correlation of immune cell infiltration (Fig 9C). The results showed the positive correlation between neutrophils cells and NK cells resting cells(r value= 0.29). T cells CD8 showed an inverse correlation with T cells CD4 memory resting (r value= -0.53). Furthermore, the bubble chart presented in Fig 9D showed that CD36 was positively correlalted with monocytes cells but negatively correlated with T cells CD4 memory resting cells (p value < 0.05).

**Fig 9.**
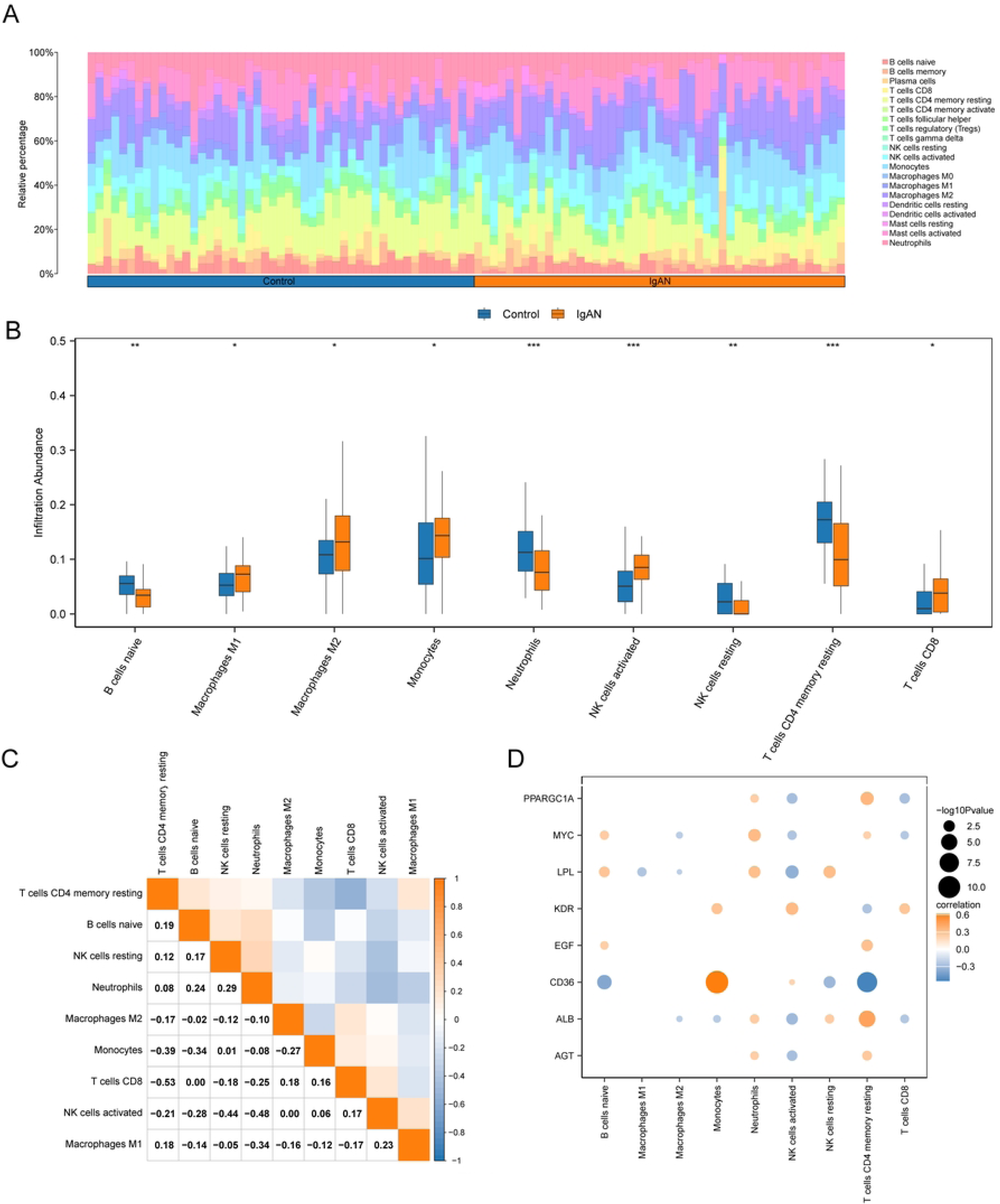
Immune infiltration analysis by CIBERSORT Algorithm in IgAN and control group. The proportion of immune cells in each sample was showed in a histogram(A) and group comparison graph (B). C: The correlation of 22 types of immune cells in IgAN renal tissues was evaluated. Red: positive correlation, blue: negative correlation. D: Bubble plot of correlation between hub genes and immune cell in IgAN. ns stands for p value ≥ 0.05, not statistically significant; * represents p value < 0.05; ** represents p value < 0.01; *** represents p value < 0.001.

### mRNA expression of hub genes in IgAN patients using nephroseq v5 online platform

The results showed that the expression levels of five hub genes (*AGT, ALB, EGF, LPL, PPARGC1A*) were significantly decreased in the glomerular tissues of IgAN patients, while that of CD36 and KDR were significantly increased compared with healthy living donors. In addition, comparing with the control group, the expression levels of MYC were significantly downregulated in the renal tubular tissues of IgAN patients(Fig 10).

**Fig 10:**
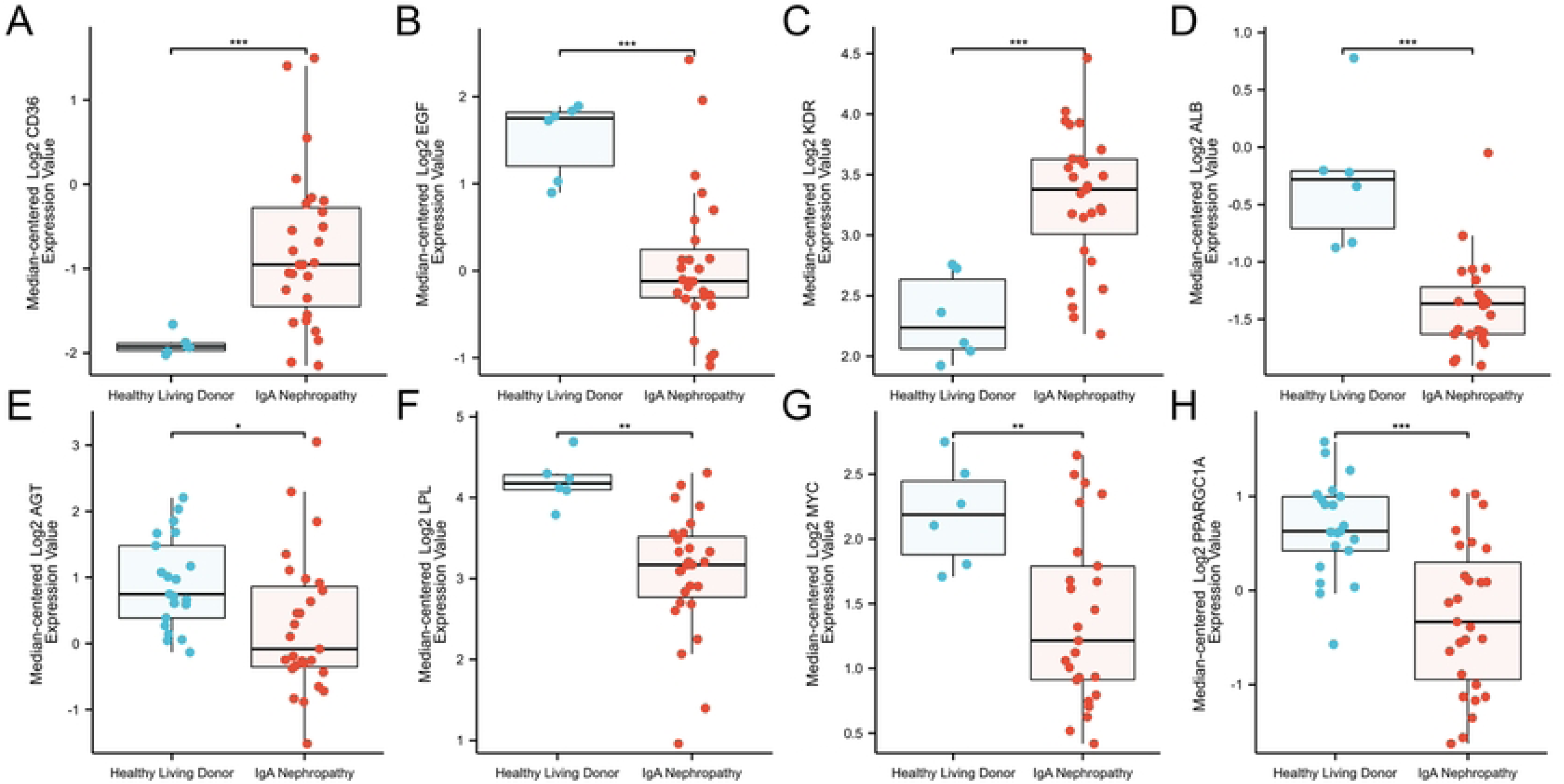
Differential expression of hub genes between IgA nephropathy and normal groups in the Nephroseq v5 database. * represents p value < 0.05; ** represents p value < 0.01; *** represents p value < 0.001.

### The validation of CD36 in IgAN

The above results revealed that CD36 has a relatively higer expression and the higher diagnositic value as well as the close correlation with immune infiltration. Based on this, CD36 was further verified in the kidney tissues of IgAN patients. The results showed that CD36 was highly expressed in the kidney tissues of IgA nephropathy patients compared to the control group(Fig 11).

**Fig 11.**
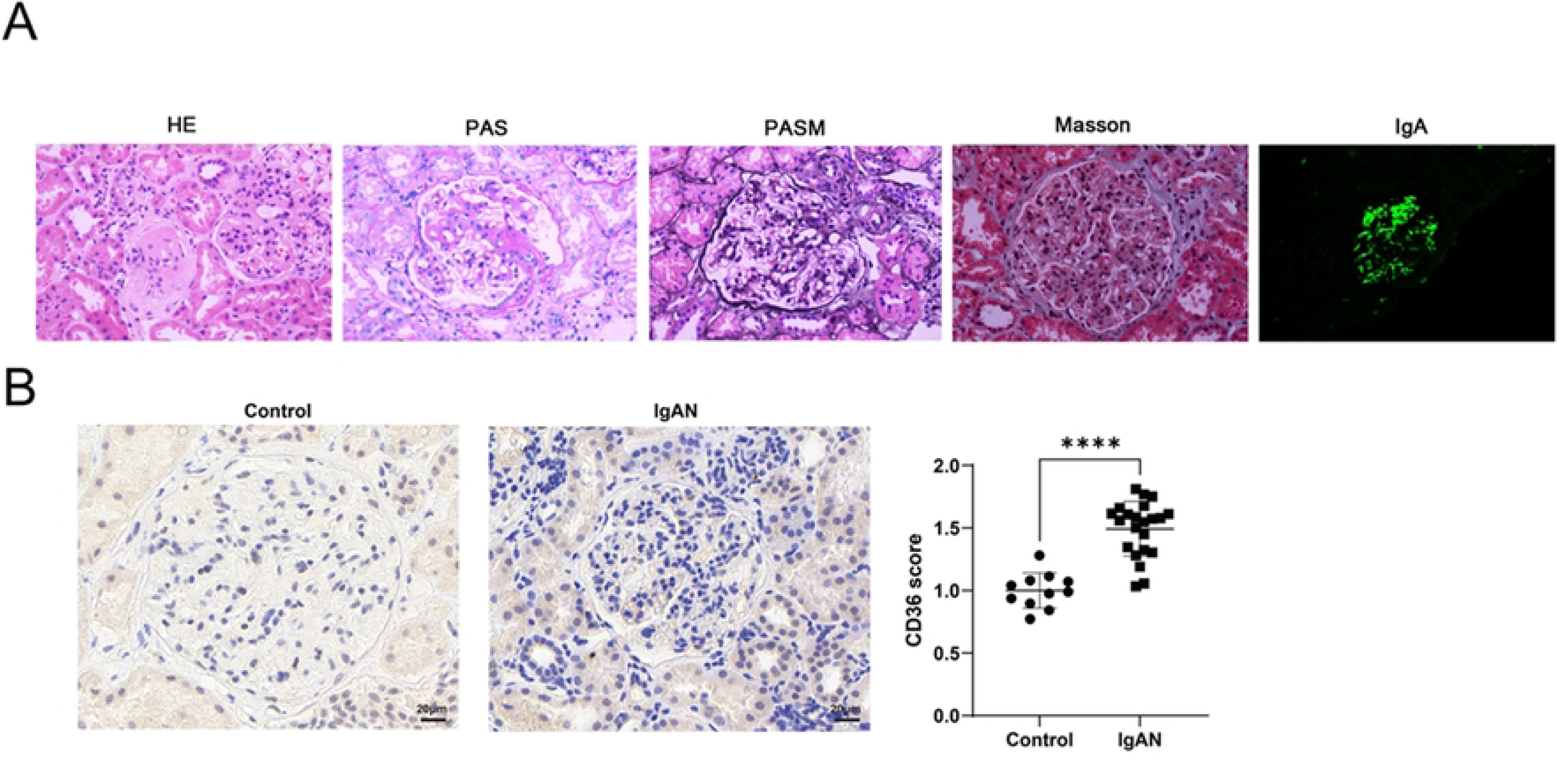
Immunohistochemistry of CD36 in IgAN renal tissues and control group. ***p < 0.001

## Discussion

The phenotypic manifestation of IgAN is highly variable, ranging from asymptomatic microscopic hematuria to rapidly progressive glomerulonephritis. The heterogeneity of clinical presentation and outcome makes it challenging to predict disease progression and treatment response. As early as 2005, Szeto et al. found chromosomal telomere of kidney cells shortened in IgAN patients by measuring DNA in the urinary sediment. Lu YY et al. reported that human IgAN was associated with increased telomere shortening and related inflammatory proteins secretion[28]. These findings suggested that telomere shortening was associated with the progression of IgAN. Although IgAN was mostly reported in young patients, emerging evidences had suggested that the incidence of IgAN was increasing in older patients, aging promote the progression of IgAN[10,28-32]. Given that shortening of telomere length was considered one of the indicators of aging, these findings suggested that aging was a risk factor for IgAN, the exact pathophysiological mechanism of aging exacerbating IgAN remains unclear. It is important to actively understand the underlying mechanisms of aging-related IgAN to develop new therapeutic strategies. Therefore, the primary purpose of the present study was to identify potential aging biomarkers in IgAN, and to explore the underlying mechanism and role of aging-related genes and immune infiltration in IgAN, so as to provide new strategy and supporting evidence for the potential mechanisms and early diagnosis of IgAN.

In this study, we identified 20 AR-DEGs in IgAN kidney samples and control groups. According to the selected AR-DRGs, GO and KEGG enrichment analysis were performed to explore potential biological functions. The results indicated regulation of macrophage derived foam cell differentiation, AGE-RAGE signaling pathway in diabetic complications, PI3K-Akt signaling pathway were related with several enriched terms, which was consistent with previous studies. Cox SN et al. reported that the genes with different expression between IgAN patients and healthy controls were mainly involved in the typical WnT-β-catenin and PI3K/Akt pathways[33]. In addition, the PI3K-Akt-mTOR pathway was also activated in IgAN podocytes, thereby reducing the level of autophagy in podocytes, leading to podocyte damage[34]. Moreover, the activation of PI3K/ Akt pathway promoted renal fibrosis and aggravated renal injury[35]. Hydroxychloroquine had been reported to protect agaist IgAN by inhibiting the PI3K/AKT signaling pathway, AGE-RAGE signaling pathway and MAPK signaling[36]. Aging may promote the accumulation of AGE, and activation of AGE-RAGE signaling pathway can enhance oxidative stress, promote the release of pro-inflammatory factors, damage vascular endothelial cells, and ultimately lead to kidney injury[37]. The lipid metabolism of macrophages consists of three different processes: cholesterol uptake, esterification and efflux. Dysregulation of these lipid metabolic pathways in turn leads to the formation of lipid-dense macrophages, which are called "foam cells." It is widely accepted that foam cells play a crucial role in the progression of atherosclerosis. Arteriosclerosis was found to be independently associated with intrarenal resistive index (RRI) in IgAN[38], and an RRI of 0.70 or higher was an independent risk factor for chronic kidney disease (CKD) progression in patients with CKD, regardless of the baseline eGFR, proteinuria, or arterial hypertension[39,40].

The GSEA results indicated that the negative regulation of notch4 signaling and inflammation response pathway were significantly enriched. Previous studies have reported that notch4 acted as a negative regulator of macrophage activation and that it was able to inhibit IFN-γ signaling, and reduce the expression of proinflammatory cytokines[41]. These inflammatory mediators promoted podocytes, renal tubule interstitium and endothelial cells injury in IgAN, which has been recognized as a chronic inflammatory disease[42]. Renal transcriptomic studies have shown that the gene expression of inflmmatory molecules were involved in the progression of renal damage in IgAN patients[43].

Subsequently, eight key genes were selected through the construction of PPI network and the selection of five algorithms: MCC, Closeness, MNC, Degree and EPC. The genes are *AGT, ALB, CD36, EGF, KDR, LPL, MYC, PPARGC1A,* respectively. Furthermore, compared with the normal control group, the expression levels of *CD36* and *KDR* were higher in IgAN patients, while the expression levels of *AGT, ALB, LPL, MYC* and *PPARGC1A* were lower. Nephroseq database also verified the differences in expression of these genes between normal population and IgAN patients. The increased expression of *CD36* was further verified on the kidney tissues of patients with IgAN by immunohistochemistry. In particular, immune cell infiltration analysis showed that *CD36* was significantly correlated with multiple immune cells. This suggested that the expression of these genes were closely related to the progression of IgAN, especially *CD36* may be a potential new therapeutic target for IgAN.

*CD36* is a transmembrane glycoprotein that belongs to a family of class B scavenger receptors. Recently, a growing number of studies had shown that *CD36* binded a variety of ligands, and thus participate in a diverse range of pathophysiological processes, such as lipid metabolism, inflammation, atherosclerosis. With the advancement of research, *CD36* play an important role in a number of kidney disease. Firstly, *CD36* overexpression led to lipid accumulation by increasing fatty acid uptake in renal tubular epithelial cells of CKD model mice and podocytes cultured in vitro[44-46]. Importantly, fatty acid accumulation and alterations in the lipid metabolism may result in glomerular and tubular cell injury[47]. In hypoxic dendritic cells, blocking *CD36* markedly reduced lipid uptake and accumulation, eliminated NKT cell overactivation, and reversed renal IRI intensification[48]. Secondly, *CD36* mediate the endocytosis and degradation of ox-LDL in macrophages. ox-LDL deposition was up-regulated in the renal tubules and interstitial compartment and correlated with fibrosis in hypercholesterolaemic mice with kidney injury following unilateral ureteral obstruction (UUO)[49]. Additionally, knockout of *CD36* gene significantly decreased oxidative stress and inflammatory pathway, thus ameliorating renal fibrosis. Compared with wild-type mice, *CD36*-deficient mice not only demonstrated reduced levels of activated NF- κ B and oxidative stress, but also decreased interstitial myofibroblasts accumulation in a hypercholesterolemic model of CKD[50,51]. In addition, *CD36* has been demonstrated to be involved in the endocytosis of many substance such as AOPP, AGEs. Previous studies indicated that binding of AOPP to *CD36* induced renal RAS activation via the PKC α -NADPH oxidase-AP-1 /NF- κ B pathway. When *CD36* was blocked, the results showed that AOPP-induced NADPH oxidase activation and ROS production were significantly inhibited[52]. Anti-*CD36* antibody treatment significantly inhibited the endocytic associatioin and degradation of AOPPs-HSA in vitro in cultured HK-2 cells. In addition to this, AOPPs endocytosis increased the production of reactive oxygen species and the secretion of transforming growth factor (TGF)- β 1 in HK-2 cells, whereas the upregulation of TGF- β 1 was neutralized in the presence of anti-*CD36* antibodies[53]. Based on the above results, it is suggested that *CD36* mediates the renal damage effect of AOPPs. AGEs accumulate has been observed during aging and age-related diseases[54]. Some studies have showed that AGEs enhance the foam cell formation and *CD36* expression in macrophages.through oxidative stress generation[55,56]. AGE-RAGE-induced oxidative stress generation stimulates ox-LDL uptake into macrophages via the Cdk5-CD36 pathway. These findings showed that CD36 play a significant role in the endocytosis of AGEs. Besides that, the expression of *CD36* and *NLRP3* were reported to be up-regulated in the podocytes of patients with lupus nephritis. Knocking out *CD36* in podocytes reduced the levels of NLRP3 inflammasome, increased the autophagy levels and alleviated podocyte injury, the present findings supported a link between autophagy and *CD36*[57]. In our study, the result showed that the expression of *CD36* was increased in the kidney tissues of IgAN compared to the control group. Consistent with this, recently, it has been reported that *CD36* was involved in the regulation of autophagy in IgAN[58]. Studies had shown that *CD36* expression was positively correlated with 24-hour proteinuria, serum creatinine, renal fibrosis-related and autophagy-related factors in IgAN. After pIgA1 stimulation, *CD36* and fibrosis-related factors in mouse mesangial cells were significantly increased. When *CD36* was knocked down, the pIgA1-stimulated mouse mesangial cells extracellular matrix accumulation was reduced. Previous studies on the mechanism of *CD36* in kidney disease were mostly seen in CKD and DKD. We speculate that *CD36* aggravate renal lipid deposition and mediate renal cell lipid peroxidation by increasing oxidative stresss and activating inflammatory response, which may contribute to the aggravation of glomerular sclerosis, podocyte injury, and promote renal fibrosis. More research is needed to further elucidate the specific molecular mechanisms in the future.

*AGT* encodes angiotensinogen, the only substrate of renin-angiotensin system (RAS) which is involved in vasoconstriction and blood pressure regulation. Blockade of the RAS is an important part of optimal supportive treatment for IgAN. In agreement with previous findings, our result showed that *AGT* gene was down-regulated in IgAN by integrated bioinformatics analysis[59,60]. There are multiple single-nucleotide variants in this gene that not only exhibit different frequency distributions among different races, but also are associated with a variety of disease risks. Previous studies have reported the expression of *AGT* in kidney is increased in patients with IgAN[61,62]. However, some studies have found no association between IgA nephropathy and *AGT*[63,64].

*ALB*, which encodes serum albumin, is not only a key protein reflecting nutritional status, but also an important regulatory inflammatory and immune response, antioxidant, transport and binding molecule[65,66]. It was reported that every 1g/L decrease in time-mean serum albumin (TA-ALB) levels, the risk of renal progression increased by 14%[67]. The study by Ni Z et al demonstrated that TA-ALB can be used to predict long-term outcomes in IgAN who have achieved remission. Consistent with the previous findings[68], we found that *ALB* was down-regulated in IgAN.

In the context of IgAN, *EGF* excretion has been reported to decrease and urinary EGF levels have been reported to predict the extent of intersitital fibrosis and renal function outcomes[69,70]. According to the Oxford classification of IgAN, urinary EGF levels were significantly negatively associated with the severity of T lesions, as well as with tubular interstitial injury[71,72]. In our study, our finding that IgAN patients had lower EGF levels than healthy individuals was consistent with previously reported results[73]. Thus, these evidences support *EGF* levels as a useful biomarker of IgA nephropathy progression.

*KDR*, which is known as vascular endothelial growth factor receptor 2 (VEGFR-2), is mainly expressed in vascular endothelial cells. As mentioned in previous studies, *KDR* plays a central role in angiogenesis by binding to VEGF, including stimulating endothelial cell proliferation, invasion, migration and survival, and increasing vascular permeability[73]. Our result showed that the expression of *KDR* was decreased in IgAN patients compared to normal group.

Lipoprotein lipase (*LPL*) is an enzyme expressed in endothelial cell membrane, which can not only hydrolyze triglycerides, but also has the function of lipoprotein uptake. Studies have shown that hypertriglyceridemia caused by *LPL* deficiency did not lead to worsen of kidney injury and atherosclerosis in a mouse model of CKD[74]. There were no significant differences in renal histology, plasma cystatin C concentration, and urine volume in mice with proximal tubular epithelial-specific *LPL* deficiency compared with control mice, suggesting that lipid abnormalities induced by *LPL* deficiency have limited effects on renal physiology or disease. A recent study showed that variants within the *LPL* gene were associated with diabetic kidney disease susceptibility and a rapid decline in kidney function in Chinese type 2 diabetes patients[75]. However, no relevant studies have been reported on *LPL* in IgAN at present.

*MYC* gene is a key member of the proto-oncogene family, which is composed of three paralogs, C-MYC, N-MYC and L-MYC. Studies have shown that it can not only regulate cell cycle progression, differentiation and apoptosis, but also but also up-regulate stress-induced transcription factor ATF4 to induce autophagy[76,77]. Our results are consistent with previous studies in which *MYC* expression is down-regulated in IgAN[60,77]. We speculate that *MYC* may be involved in the progression of IgAN through autophagy mechanisms. In addition, it has been reported that in familial IgAN, *MYC* is a key gene modulator of the WNT/ β -catenin and PI3K/ AKT pathways, which are also one of the pathogenesis of IgAN[78].

Multiple studies have shown that peroxisome proliferator-activated receptor γ coactivator-1 α (*PGC-1 α* or *PPARGC1A*), as a major regulator of mitochondrial biogenesis and energy metabolism, plays a crucial role in a variety of acute and chronic kidney diseases[79]. The expression of *PGC-1 α* gene was significantly decreased in CKD patients, and the expression level is significantly negatively correlated with interstitial fibrosis and positively correlated with glomerular filtration rate[80]. Our results suggested that *PGC-1α* expression was also down-regulated in IgAN. However, *PGC-1 α* expression and its effects have not yet been reported in IgAN at present, and further studies are needed in the future.

There is growing evidence suggesting the crucial role of immune system in the pathogenesis of IgAN. Our immunoinfiltration analysis revealed significant differences in 9 types of immune cells between IgAN patients and controls, suggesting a specific immune landscape associated with the disease. Our results indicated a decreased infiltration of macrophages M1, macrophages M2, monocytes, NK cells activated, T cells CD8, along with an increased infiltration of B cells naive, T cells CD4 memory resting, NK cells resting, neutrophils, suggesting that these immune cells may be involved in the pathogenesis of IgAN. It is worth noting that the number and subtypes of macrophages were correlated with the clinical manifestations and pathology of IgAN patients, and they may contribute to renal sclerosis, glomerular injury and renal fibrosis through the production of pro-inflammatory cytokines, growth and fibrotic factors[81,82]. T cells are another pivotal component of the adaptive immune response in IgAN. Many studies have confirmed that most T cell subpopulations were associated with the clinical features of IgAN[83,84]. Their involvement is also noteworthy given the role of B cells in IgA production and immune complex formation, both of which are central features of IgAN pathogenesis. In addition, studies have confirmed that immunomodulators targeting B cells can significantly improve IgAN, regulate the secretion of cytokines and chemokines, and delay the progression of the disease[85,86]. *CD36* is associated with multiple immune cell subpopulations in kidney tissue of IgAN patients, suggesting that *CD36* not only serve as a potential biomarker for diagnosis, but also offer a window into understanding how genetic regulation intersects with immune processes to drive disease progression.

Despite the promising findings of our research, there are several limitations that may impact the interpretation and applicability of our results. Firstly, this study was based on the previously published data set for secondary utilization and analysis. Secondly, the sample size of the data set was not large. Further experimental and clinical studies are required to validate the results and clarify the molecular mechanisms underlying the observed associations.

## Conclusions

In conclusion, 20 AR-DEGs were identified in IgAN from the perspective of bioinformatics analysis, which were mainly enriched in regulation of macrophage derived foam cell differentiation, AGE-RAGE signaling pathway in diabetic complications, PI3K-Akt signaling pathway. Our study also suggested a correlation between the expression levels of these genes and the proportion of infiltrating immune cells in patients with IgAN. *CD36* has certain diagnostic value for age-related IgAN and can be used as a potential biomarker for age-related IgAN.

## Data Availability

All relevant data are within the manuscript and its Supporting Information files.

## Acknowledgements

We acknowledge GEO database for providing their platforms and contributors for uploading their meaningful datasets.

## Supporting information

**S1 Table. Table 1 GEO Microarray Chip Information.**

**S2 Table. Table 2 Results of GSEA for Combined Datasets.**

**S3 Table. Table 3 Results of GO and KEGG Enrichment Analysis for ARDEGs**

